# Epidemiological investigation of the first 135 COVID-19 cases in Brunei: Implications for surveillance, control, and travel restrictions

**DOI:** 10.1101/2020.06.29.20142463

**Authors:** Justin Wong, Liling Chaw, Wee Chian Koh, Mohammad Fathi Alikhan, Sirajul Adli Jamaludin, POH Wan Wen Patricia, Lin Naing

## Abstract

**Background:** Studies on the early introduction of SARS-CoV-2 in a naïve population have important epidemic control implications. We report findings from the epidemiological investigation of the initial 135 COVID-19 cases in Brunei and describe the impact of control measures and travel restrictions.

**Methods:** Epidemiological and clinical information were obtained for all confirmed COVID-19 cases in Brunei, whose symptom onset was from March 9 to April 5, 2020 (covering the initial 5 weeks of the epidemic). Transmission-related measures such as reproduction number (R), incubation period, serial interval were estimated. Time-varying R was calculated to assess the effectiveness of control measures.

**Results:** A total of 135 cases were detected, of which 53 (39.3%) were imported. The median age was 36 years (range = 0.5 to 72). 41 (30.4%) and 13 (9.6%) were presymptomatic and asymptomatic cases respectively. The median incubation period was 5 days (IQR = 5, range = 1 to 11), and the mean serial interval was 5.39 days (sd = 4.47; 95% CI: 4.25, 6.53). R0 was between 3.88 and 5.96, and the doubling time was 1.3 days. By the 13th day of the epidemic, the Rt was under one (Rt = 0.91; 95% credible interval: 0.62, 1.32) and the epidemic was under control.

**Conclusion:** Epidemic control was achieved through a combination of public health measures, with emphasis on a test-isolate-trace approach supplemented by travel restrictions and moderate physical distancing measures but no actual lockdown. To maintain suppression, regular and ongoing testing of high-risk groups can supplement the existing surveillance program.

## Introduction

The global spread of COVID-19 and the lack of an effective vaccine or therapeutic options pose challenges for disease control^1^ and travel health.^2^ Importation events in a country with no cases can lead to an exponential increase in case numbers within a short time period.^3^ As such, countries have implemented travel restrictions in response to the global rise, however, its effectiveness is debatable.^4, 5^ Studies on the early introduction of the virus in a naïve population can provide insight into the natural history of the disease and have implications for control measures.

Brunei Darussalam (pop. 459,500), a well-connected country vulnerable to multiple importation events, detected its first COVID-19 imported case on March 9, linked to an international super spreading event (SSE), the Sri Petaling Mosque Tablighi Jamaat cluster, previously described.^6, 7^ Given the absence of widespread community transmission, and slowing trajectory since the 100th confirmed case, Brunei’s response to this first wave of the pandemic has been generally successful.^8^ To maintain this, a number of questions need to be addressed. First, what are the epidemiological characteristics of the cases observed so far? Second, what is the role of travel-related cases in driving the disease? Third, how infectious are the cases and how effective are approaches to reducing transmission?

Here, we report findings from the first 135 COVID-19 cases, detected within the first 5 epidemiological weeks of the local epidemic, along with their epidemiological, clinical and transmission characteristics. As jurisdictions that have implemented lockdowns begin to bring the epidemic under control,^9^ our findings will be important to calibrate detection and response efforts in potential future waves of the pandemic.

## Methods

### Case identification and contact tracing

The surveillance and contact tracing strategy has previously been described.^7^ Since January 23, clinical and laboratory surveillance has been implemented across the country, and the testing criteria has progressively expanded in scope (**Supplementary Table 1**). A confirmed case is someone who tested positive for SARS-CoV-2 through real-time reverse transcriptase polymerase chain reaction (RT-PCR) test on nasopharyngeal (NP) swab. All laboratory-confirmed COVID-19 cases with symptom onset from March 9 to April 5, 2020 were included in this study and followed-up until recovery or death.

Epidemiological investigation is conducted for each confirmed case, and information is collected on demographic characteristics, clinical symptoms, travel history, activity mapping two days prior to onset of symptoms (or swab date for asymptomatic cases), and contact history. A close contact is any person living in the same household, or someone within one meter of a confirmed case in an enclosed space for more than 15 minutes. All close contacts undergo RT-PCR testing. Those who tested negative were quarantined at home for 14 days from last exposure, and those who later developed symptoms were re-tested. All confirmed COVID-19 cases are treated and isolated at the National Isolation Centre (NIC). Cases were discharged following two consecutive negative SARS-CoV-2 NP swabs collected at ≥ 24-hour intervals.

We categorised cases into two: imported cases (defined as individuals presumed to have acquired the infection outside Brunei), and locally transmitted cases (defined as those without travel history).

### Statistical analysis

An epidemic curve was constructed based on the date of symptom onset (for symptomatic and presymptomatic cases) and the date of NP swab collection (for asymptomatic cases). Duration between symptom onset date to diagnosis date was calculated. Exposure period between imported cases and their close contacts was calculated as the duration between their return date to Brunei and their diagnosis/swab collection date. Incubation period was calculated as the duration between the known exposure date of confirmed cases and their symptom onset dates. Serial interval (SI) was calculated as the duration between the symptom onset date for the primary case and that of secondary case. Only symptomatic or presymptomatic infector-infectee pairs with clear epidemiological links were included in the SI calculation. Group comparison was done between the imported and local contact cases, using Chi-square, Fisher’s Exact or Mann-Whitney’s tests as appropriate.

The estimation of the reproduction number (defined as the expected number of secondary cases infected by a primary case) was done using two methods. The basic reproduction number (R0) was estimated from the mean SI and the exponential growth rate of the cumulative number of cases in the epidemic 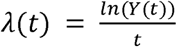, using the formula R0 = 1 + SIλ + *f* (1-*f*)(SIλ)^2^, where *f* is the ratio of the infectious period to the SI. This assumes an exponential distribution, allowing for a range of reported values.^10^ The exponential growth phase of between March 5 and 10 was chosen for the R0 estimation because it represents the initial growth of the curve (**see Figure 1**), and also because control measures were enhanced after March 9. This 6-day period represents more accurately the true nature of SARS-CoV-2 transmission, in the absence of heightened measures. The epidemic growth rate and doubling time was also calculated, based on established formulae.^11^

**Figure 1.**
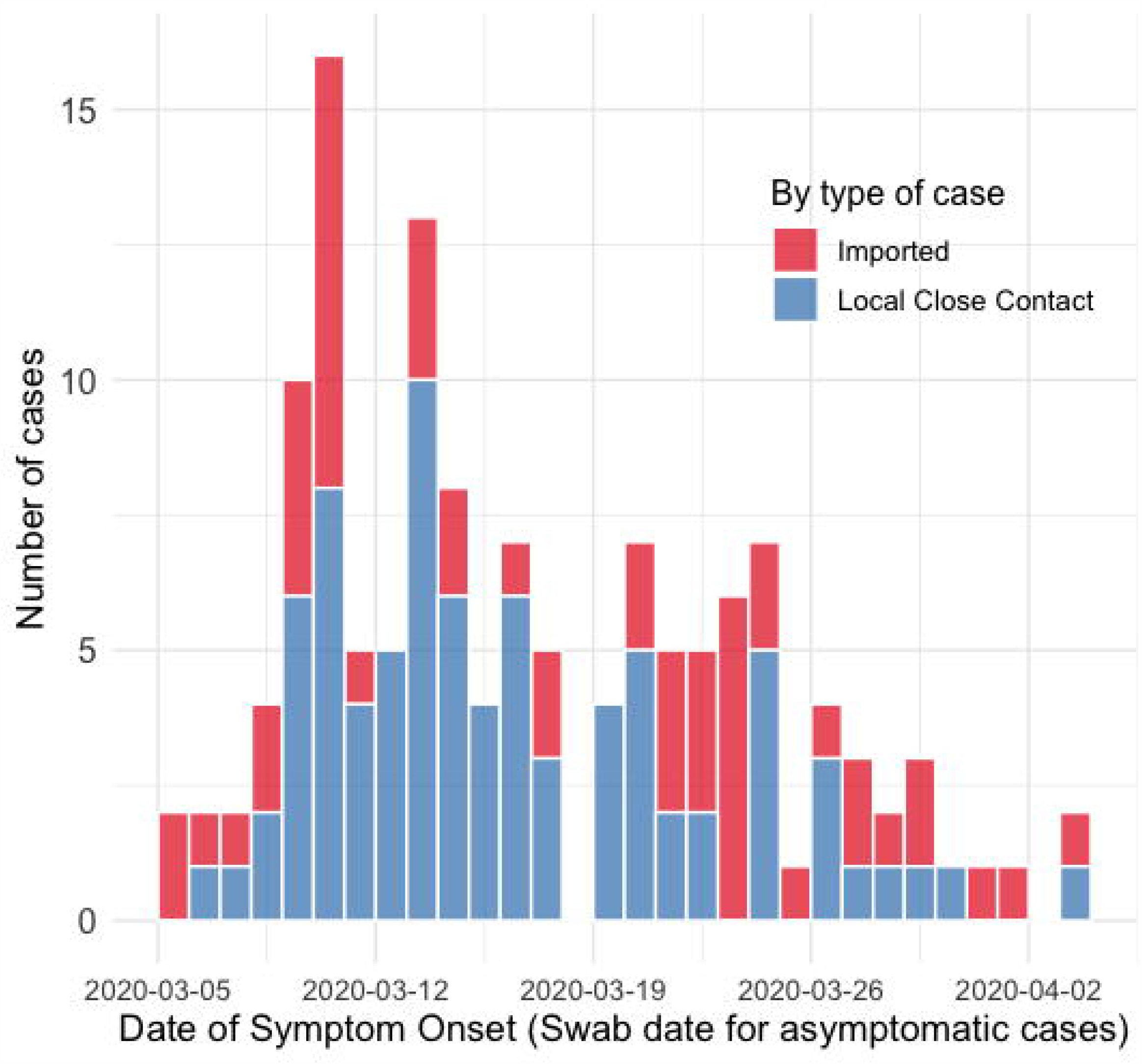
Epidemic curve for the first 135 COVID-19 cases in Brunei Darussalam, by imported (red bars) and locally transmitted cases (blue bars).

We also estimated the time-varying reproduction number (Rt) to assess the effectiveness of epidemic control measures, based on methods proposed by Thompson et al.^12^ and using the EpiEstim 2.2 package. This Rt estimation uses case incidence data and SI distribution, and distinguishes between imported and locally transmitted cases. Following studies that use a non-negative SI distribution to approximate the distribution of the generation time,^13, 14^ we used a Gamma distribution and incorporated uncertainty in the parameters (sd of mean = 1, sd of standard deviation = 0.5). The median Rt and 95% credible intervals for each day were estimated jointly from incidence data and from the posterior SI distribution, using a 6-day sliding window. This 6-day window reduces the bias associated with early estimations of Rt, as at least one average SI has passed.^15^

All analyses were conducted using Microsoft Excel and R (ver. 3.6.3).^16^ A p-value < 0.05 was considered as statistically significant. Ethics approval was granted by the University Research Ethics Committee, Universiti Brunei Darussalam (Ref: UBD/OAVCR/UREC/Apr2020-05).

## Results

### Description of cases

A total of 135 cases were detected in the first 5 weeks, comprising 53 (39.3%) imported and 82 (60.7%) locally transmitted cases **(Table 1)**. The median age was 36 years (ranging from 6 months to 72 years). 53 (39.3%) cases were female. When compared to imported cases, locally transmitted cases were significantly younger (p= 0.002) and had more females (p= 0.008).

**Table 1:** Demographic and clinical characteristics of the first 135 COVID-19 cases in Brunei

|  |  | **All cases**<br>**(n=135)**<br>**n(%)** | **Imported**<br>**cases (n=53)**<br>**n(%)** | **Local contact**<br>**cases (n=82)**<br>**n(%)** | **p-value** |
| --- | --- | --- | --- | --- | --- |
| **Median age in years**<br>**(IQR; min. to max.)** |  | 36.0<br>(27; 0.5 to 72) | 39.0<br>(27; 17 to 68) | 31.5<br>(26; 0.5 to 72) | **0.002** |
| **Age group** | 0-9 | 7 (5.2) | 0 (0.0) | 7 (8.5) | **< 0.001** |
|  | 10-19 | 16 (11.9) | 3 (5.7) | 13 (15.9) |  |
|  | 20-29 | 26 (19.3) | 7 (13.2) | 19 (23.2) |  |
|  | 30-39 | 28 (20.7) | 17 (32.1) | 11 (13.4) |  |
|  | 40-49 | 18 (13.3) | 4 (7.5) | 14 (17.1) |  |
|  | 50-59 | 23 (17.0) | 10 (18.9) | 13 (15.9) |  |
|  | 60-69 | 17 (12.6) | 12 (22.6) | 5 (6.1) |  |
| **Gender** | Female | 53 (39.3) | 13 (24.5) | 40 (48.8) | **0.008** |
|  | Male | 82 (60.7) | 40 (75.5) | 42 (51.2) |  |
| **Comorbidity** | Obesity | 6 (4.4) | 3 (5.7) | 3 (3.7) | 0.679 |
|  | Heart disease | 6 (4.4) | 3 (5.7) | 3 (3.7) | 0.679 |
|  | Respiratory disease | 7 (5.2) | 2 (3.8) | 5 (6.1) | 0.704 |
|  | Diabetes mellitus | 7 (5.2) | 5 (9.4) | 2 (2.4) | 0.111 |
|  | Hypertension | 18 (13.3) | 11 (20.8) | 7 (8.5) | 0.067 |
|  | Hyperlipidaemia | 18 (13.3) | 12 (22.6) | 6 (7.3) | **0.018** |
| **Symptom status** | Symptomatic | 81 (60.0) | 29 (54.7) | 52 (63.4) | 0.596 |
|  | Presymptomatic | 41 (30.4) | 18 (34.0) | 23 (28.0) |  |
|  | Asymptomatic | 13 (9.6) | 6 (11.3) | 7 (8.5) |  |
| **Duration between**<br>**symptom onset (or**<br>**swab taken) and**<br>**diagnosis<sup>^</sup>** | -8 to 0 days | 44 (32.6) | 17 (32.1) | 27 (32.9) | 0.665 |
|  | 1 to 2 days | 39 (28.9) | 18 (34.0) | 21 (25.6) |  |
|  | 3 to 5 days | 26 (19.3) | 8 (15.1) | 18 (22.0) |  |
|  | > 5 days | 26 (19.3) | 10 (18.9) | 16 (19.5) |  |
| **Severity<sup>#</sup>** | Asymptomatic | 13 (9.6) | 7 (13.2) | 6 (7.3) | 0.352 |
|  | Mild | 101 (74.8) | 36 (67.9) | 65 (79.3) |  |
|  | Moderate | 14 (10.4) | 7 (13.2) | 7 (8.5) |  |
|  | Severe | 2 (1.5) | 0 (0.0) | 2 (2.4) |  |
|  | Critical | 5 (3.7) | 3 (5.7) | 2 (2.4) |  |

\*Cases were classified as: (i) symptomatic, if symptoms were reported on or prior to NP swab collection day, (ii) presymptomatic, if symptoms were reported after NP swab sample was taken but during admission, and (iii) asymptomatic, if no symptoms were reported since NP swab collection day until date of hospital discharge.
<sup>^</sup>This includes the asymptomatic cases, from whom the symptom onset date was replaced by date of swab collection.
<sup>#</sup>Severity was classified as (i) asymptomatic, for those with no symptom throughout their disease; (ii) mild, for patients who had uncomplicated upper respiratory tract infection symptoms and no radiological changes; (iii) moderate, for patients with radiological changes but did not require supplemental oxygen; (iv) severe, for patients who showed signs of severe pneumonia including tachypnoea > 30/minute, SpO<sub>2</sub> of ≤ 93% on room air or abnormal arterial blood gases, as well as patients showing signs of sepsis with evidence of organ dysfunction; and (v) critical, for patients who developed
septic shock, i.e. persistent hypotension requiring vasopressors support to maintain mean arterial pressure $\geq 65$ and those who developed acute respiratory distress syndrome requiring ventilatory support.

81 (60%) of cases developed symptoms, reported either during or prior to NP sample collection. Notably, we observed high proportions of presymptomatic and asymptomatic cases - 41 (30.4%) and 13 (9.6%) respectively. 61.5% of the cases were detected within 2 days of symptom onset or NP swab date. Among them, 32.6% (n= 44) were detected on or before the day of symptom onset. No significant differences were observed between imported and locally transmitted cases **(Table 1)**. The most common reported symptoms were fever (62.2%), sore throat (62.2%) and cough (59.3%) **(Supplementary Table 2)**.

Among these 135 cases, 3 subsequently died from COVID-19 complications, giving a case fatality rate of 2.2%. All 3 deaths were in men, aged 64, 56, and 67 years, respectively.

### Impact of travel restrictions

Since late-January, travel restrictions have been progressively implemented in response to the emerging regional and later global situation. Initially, travellers from Hubei province, China were restricted, while those from other parts of mainland China underwent 14 days home quarantine. These restrictions were gradually tightened, to first of all restricting travellers from Iran and Italy (the then emerging epicentres) and mandating quarantine for travellers from China and South Korea. These restrictions had a considerable impact on arrivals into Brunei. Immigration data indicate a 20.9% decrease in arrivals from January to March 2020 (872,315 people) compared to the same period in 2019 (1,103,028 people).

Outbound travel was restricted for all Brunei residents on March 15, and a ban on all foreign citizens entering the country was enacted on March 23. All individuals entering Brunei from March 20 underwent 14-day quarantine at a designated facility, and RT-PCR testing on arrival.

The origin country of imported cases changed over time – initially Malaysia (20 cases), and as the epidemic progressed globally, and overseas Brunei citizens were returning, imported cases were identified in travellers and returning residents from Indonesia (n=14), United Kingdom (n=11), Thailand (n=2), USA (n=2), Austria (n=1), Cambodia (n=1), Australia (n=1), and the Philippines (n=1).

**Figure 1** shows the epidemic curve, by the date of symptom onset or NP swab date for asymptomatic cases. Detection of positive cases among local close contacts occurred very early in the epidemic. Coupled with the early detection of further generations **(Supplementary Figure 1)**, this suggests a short time interval for transmission within the community.

### Epidemic characteristics over time

The mean duration from onset to diagnosis for local transmitted cases decreased from 9 days in the first week to −1.7 days in the fifth week of the epidemic **(Fig 2A)**. Among imported cases, this reduction occurred between the third and fourth week of the epidemic (from 7.3 to 1.3 days, respectively), coinciding with implementation of quarantine and testing on for all arrivals on March 20 **(Fig 2A)** demonstrating the impact of increasingly stringent travel restrictions. There were no local infections linked to those imported cases in the last two weeks of the epidemic.

**Figure 2.**
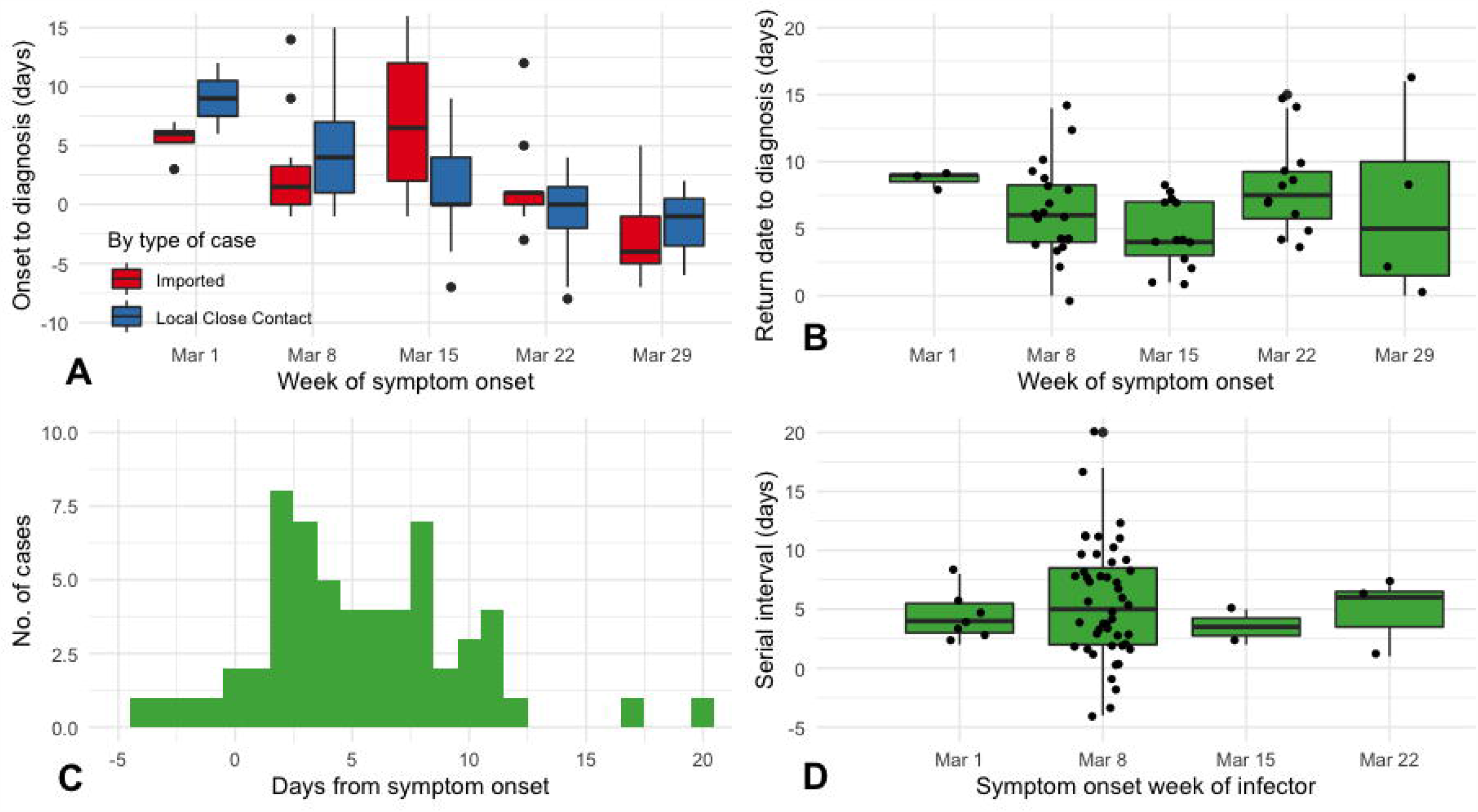
**A (top left)** Boxplot of the duration between symptom onset to diagnosis among symptomatic and presymptomatic cases only, by week of symptom onset and by type of case: imported (in blue) and local case contact (in red). Black dots above and below each boxplot indicate outliers; **B (top right)** Boxplot of the exposure period of the 53 imported cases to their local contact cases (using their return dates to Brunei as start point), by week of symptom onset. Black dots represent each case, jittered for visual clarity; **C (bottom left)** Histogram showing the serial interval (SI) distribution for the 59 symptomatic infector-infectee pairs; **D (bottom right)** Boxplot showing variations in the SI distribution by the symptom onset week of the infector. Black dots represent each case, jittered for visual clarity.

Using the known return dates from imported cases (n= 53), the median duration in the exposure period to other contacts was 7 days (IQR = 5), ranging between 1 and 16 days. The reduction in the exposure period between imported cases and their close contacts from their return to diagnosis was apparent between the third (7.7 days) and fourth week (4.2 days; **Fig 2B)**. Summary statistics and distribution remained unchanged when the 15 local contact cases with known exposure dates were included. Among 82 local contact cases, 15 of them (18.3%) had known exposure dates to confirmed imported cases. Using these dates, the median incubation period is 5.0 days (IQR = 5), ranging between 1 and 11 days.

Based on 59 symptomatic and presymptomatic infector-infectee pairs, the mean SI was 5.39 days [sd = 4.47; 95% CI: 4.25, 6.53 (approximated using normal distribution)]. The range for the serial interval was between −4 to 20 days **(Fig 2C)**. Four pairs (6.8%) had negative SI values. The mean SI was relatively constant throughout the 4 weeks of the epidemic **(Fig 2D)**.

Using the calculated mean SI and the 6-day growth phase of the epidemic, the growth rate was 0.54/day and the R0 in the early phase of the epidemic ranges between 3.88 and 5.96. The doubling time was 1.3 days.

**Figure 3** shows the estimated Rt and the timing of the control measures implemented after March 11. The initial median reproduction number was estimated to be 2.19 (95% credible intervals: 0.86, 4.99) on the 7th day of the epidemic (March 11). Rt gradually decreased after several control measures were put in place and was below one on the 13th day (March 17) and reached 0.20 (95% credible intervals: 0.06, 0.49) on the 30th day (April 3).

**Figure 3.**
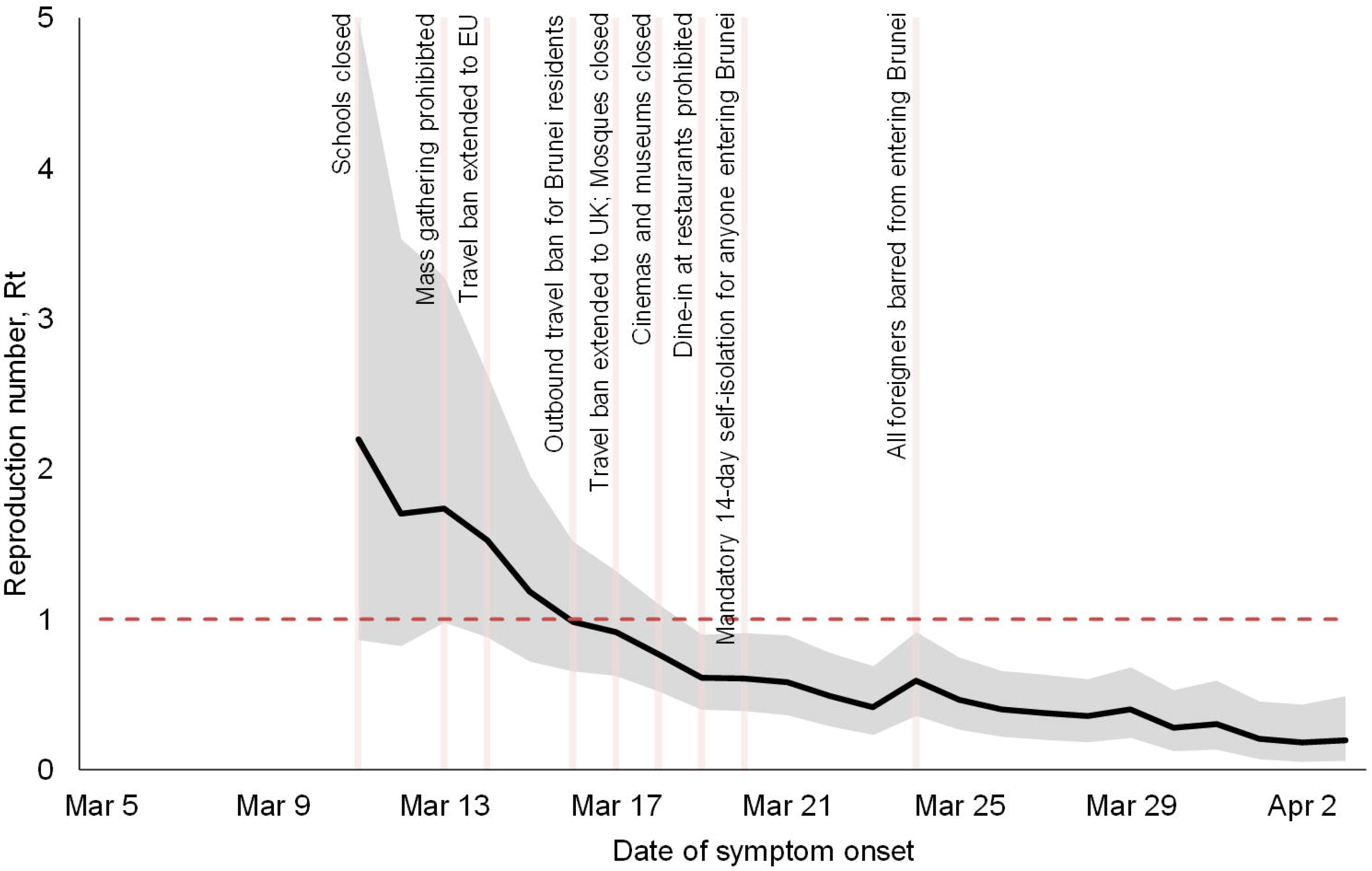
Time-varying reproduction number (Rt) estimates of the COVID-19 epidemic in Brunei. A 6-day sliding window was used. The black solid line is the estimated median R_t_ and the grey areas are the 95% credible intervals.

However, because these control measures were implemented quickly over a short time period, it is difficult to attribute the observed reduction in Rt to a specific intervention.

## Discussion

### Key findings

A total of 135 cases (53 imported and 82 locally transmitted) were reported from the start of the epidemic on March 5 up till the first week of April. All locally transmitted cases could be traced to an importation event, and there were no cases detected without a clear epidemiological link. Brunei has managed to successfully control the first wave of the COVID-19 pandemic. The R0 was between 3.88 and 5.96, and the epidemic had a doubling time of 1.3 days during the exponential phase. There was a rapid decline such that by day 13, the Rt was under 1.

### Estimating the reproduction number

Assuming the ratio of the infectious period to the serial interval (*f*) is 0.3, the R0 is 5.63, higher than those estimated by WHO (1.9 – 2.5) however, it is within those estimated in other studies from China^17-19^ and Europe.^20^ Our observations of negative SIs pose challenges for specifying *f*. Hence we reported R0 as a range covering all plausible values of *f*, using the mean SI value from our data and with assumption of exponential growth. Then, Rt was estimated from the 7th to 30th day. The 95% credible intervals of the initial Rt value (0.86 to 4.99) are consistent with the R0 estimate.

### Test, isolate and trace

We observed that the approach of isolation of confirmed cases, as well as tracing and quarantine of their contacts, on its own, resulted in significantly reducing the effective reproduction number. By the time the early physical distancing measures had been implemented, the Rt had reduced from 5.63 (ranging between 3.88 and 5.96) at the start to 2.19 (95% credible bands: 0.86, 4.99) by the 7th day (March 11). Our findings on the importance of the test, isolate, and trace approach concord with observational studies from Hong Kong^21^ and Singapore.^22^

We highlight three characteristics of this approach as implemented in Brunei. First, testing was conducted on a significant scale. Even prior to detection of the first case, testing was already conducted for all inpatient pneumonia cases, in addition to those who met the suspect case definition. 24-hour testing centres were established within 2 days of the detection of the first case (March 11), and all contacts were tested regardless of symptom status.^8^ The relatively high proportion of asymptomatic (9.6%) and pre-symptomatic (30.4%) cases identified demonstrate both the breadth of testing, and the speed at which cases were identified.

Second, all confirmed cases (regardless of disease severity and symptom status) were isolated in a dedicated isolation facility and remained isolated until two negative results were obtained from RT-PCR specimens at 24-hour intervals. This reduced the chance of household transmission which could not be excluded if cases were allowed to isolate at home.^23^ Finally, contact tracing was conducted for each case, using a variety of tools including case interview, workplace assessment, and mobile phone data. Contacts were placed on 14-day home quarantine and in-person spot checks with penalties for non-compliance were also conducted.

### Travel restrictions and other non-pharmaceutical interventions

We report several characteristics of SARS-CoV-2 that make effective isolation and contact tracing challenging, including high transmissibility, a relatively short SI (mean SI = 5.39 days), and a high proportion of asymptomatic and presymptomatic cases suggesting the potential for silent transmission. As such, the test, isolate, trace approach was supplemented with physical distancing measures to increase the likelihood of achieving sustained control.^24^

Restricting travel is one measure by which countries have responded to the COVID-19 pandemic.^25^ By the time Brunei implemented an exit travel ban and restricted the entry of foreign citizens in the country, the Rt was already decreasing and had neared 1. Nonetheless, we suggest that reducing ongoing vulnerabilities to importation events, through restrictions on incoming travelers, and requirements for testing and quarantine for all arrivals in the country limited additional spread as observed in the reduction in mean time from symptom onset to diagnosis observed for imported cases following implementation of mandatory quarantine and testing for all arrivals. Modelling studies suggest a role for travel restrictions in containing the epidemic, with one model estimating that travel restrictions in Wuhan reduced case importations elsewhere by nearly 80% until mid-February.^26^

While various other physical distancing measures were implemented, including school closures, prohibition on mass gatherings, cinemas, and religious services, and dine-in restrictions, importantly no lockdown was implemented and there were no generalised stay-at-home orders. Most businesses and government agencies were able to operate. These suggest that effective test, isolate, and trace approaches were able to control the epidemic with moderate levels of physical distancing.^27^ This finding corroborates the experience of other countries. In Hong Kong, case isolation and contact tracing were combined with other physical distancing measures (but no lockdown), which resulted in an estimated effective reproduction number near 1 for 8 weeks.^28^ In South Korea, testing and tracing has been combined with school closures and remote working.^29^

Even with the best efforts at testing, case identification, and quarantine, the potential for widespread community transmission is clear. Once established, suppression may necessitate the implementation of severely disruptive social distancing measures.^30, 31^

### Limitations

Our study has several limitations. First, while we can be reasonably confident of having identified most cases since March, given more restrictive testing criteria in January and February, we are unable to account for potential importation events that may have occurred prior to detection of the first case. Second, the generalizability of our results are limited due to a lack of cases in settings such as residential care facilities and dormitories, no community transmission, and small number of cases. Third, due to the potential for pre-symptomatic infection of SARS-CoV-2, using the SI distribution to approximate the generation time distribution is problematic. We had not accounted for negative SIs with the use of the Gamma distribution and could have overestimated Rt to fit incidence data. One way to account for negative SI is to use a deconvolution approach using the incubation period distribution to recover the generation time distribution.^32^ However, this assumes that the generation time and incubation period distributions are independent, which may not be appropriate. Finally, given the limited data available and the analytic methods employed, we could not directly estimate the effectiveness of other NPIs, such as face-mask wearing, personal hygiene practices, and voluntary reductions in mobility.

## Conclusion

Swift control of COVID-19 in Brunei was achieved through a combination of public health measures, focusing on a test-isolate-trace approach supplemented by travel restrictions and general physical distancing measures but no actual lockdown. Looking forward, to ensure suppression is maintained, regular and ongoing testing of high-risk groups such as those working in residential institutions, health care workers, and those at greater the risk of severe complications can supplement the existing surveillance program. Additionally, leveraging technological tools that speed up the process of contact tracing, and timely re-imposition of physical distancing measures if necessary, can reduce the likelihood of seeding a second wave in Brunei.

## Data Availability

Data is available from the corresponding author, upon reasonable request.

